# Population-Based Prevalence of Overweight and Obesity in German Youth: Findings from MoMo 2.0 and Comparison with KiGGS

**DOI:** 10.64898/2026.10.01.26364513

**Authors:** Claudia Lazarides, Ronny Kuhnert, Anja Schienkiewitz, Lara Tschuschke, Leon Klos, Janis Fiedler, Laura Wolbring, Anke Hanssen-Doose, Carmen Volk, Claudia Niessner, Julika Loss, Manuel C. Voelkle, Alexander Woll

## Abstract

**Objective:** Overweight and obesity in childhood and adolescence are major risk factors for health problems across the lifespan. This manuscript reports the current prevalence of overweight and obesity among children and adolescents aged 4-17□years living in Germany and compares them with previous population-based results.

**Methods:** A representative, cross-sectional sample of 4-17-year-olds was drawn following a multistage sampling process. Analyses are based on weight and height of 3,978 individuals measured in the field by trained test instructors between 2023-2025. Overweight and obesity classification was defined using age- and sex-specific body-mass-index percentiles. To allow for comparability of prevalence estimates derived from previous assessment waves and other panel studies dating back to 2005, an alignment factor was derived using an inverse probability of participation weighting approach. Together with sampling weights, the alignment factor was applied to obtain current, representative prevalence estimates of overweight and obesity.

**Results:** The aligned, weighted prevalence of overweight and obesity was 16.1% (95%□CI□[14.5-17.9%]), whereas prevalence of obesity was 6.7% (95%□CI [5.5-8.1%]). Age- and sex-specific prevalence rates are reported and compared to previous waves.

**Conclusions:** The current prevalence of overweight and obesity in children and adolescents aged 4-17 years living in Germany remains stable over the past two decades.

**Study Importance Questions:** What is already known about this subject?

- Youth overweight and obesity are a major public-health problem with lasting health consequences
- The last national German estimates (KiGGS, 2014-2017) are nearly a decade old, and newer studies are not directly comparable.

What are the new findings in your manuscript?

- In 2023-2025, 16.1% of German 4- to 17-year-olds were overweight and 6.7% had obesity, compared to KiGGS stabilizing over the last 20 years
- Modern alignment methods allow valid prevalence estimation across study designs and sample procedures, in the absence of new health surveillance data

How might your results change the direction of research or the focus of clinical practice?

- The plateau is no reason for complacency: effective structural prevention policies still need to be implemented and evaluated, continued health monitoring is necessary
- Future research should examine socioeconomic subgroups and use aligned surveillance to track whether the plateau persists.

## Introduction

Overweight and obesity in childhood and adolescence constitute a major global public⍰health problem.(1) Excess weight in early life persists into adulthood (2) and is associated with adverse health outcomes, such as an increased risk of cardiometabolic diseases (3) and psychosocial impairments (4) and imposes substantial societal costs.(5) To reduce the adverse health consequences of overweight and obesity among children and adolescents, continuous, population-representative monitoring of overweight and obesity is needed to inform the planning and evaluation of preventive measures and interventions.(6, 7)

Nearly a decade has passed since nationally representative estimates of excess weight among youth in Germany were last updated limiting the assessment of temporal trends. The most recent estimates based on national reference systems were derived from wave 2 of the German Health Interview and Examination Survey for Children and Adolescents (KiGGS; 2014-2017), which reported a prevalence of 15.4% for overweight and 5.9% for obesity among 3- to 17-year-olds.(8) In the absence of more recent KiGGS data, updated estimates must draw on data from other population-based surveillance studies; however, differences in study design and sample composition may limit direct comparability.

Modern alignment methods offer a way to link such surveillance data to established population studies and thereby generate methodologically comparable estimates.(9, 10) In this study, we apply an alignment approach to data from the Motorik-Module (MoMo) 2.0 study to provide current estimates of overweight and obesity among children and adolescents in Germany that are comparable with previous KiGGS estimates.

## Methods

### Study Design

KiGGS (11) monitored overweight and obesity in youth aged 3-17 years in the baseline assessment (2003-2006) and wave 2 (2014-2017) as part of the health monitoring system at the Robert Koch Institute. The Motorik-Modul Study (MoMo) supplemented KiGGS with in⍰depth investigations of motor performance and physical activity in youth aged 4-17 years. MoMo participants were recruited out of the entire KiGGS sample in both waves. The MoMo 2.0 Study (MoMo 2.0, 2023-2025; 12) established a new population-based sample including anthropometric data. The three assessment waves (2003-2006, 2014-2017, 2023-2025) collected data in a cohort⍰sequential (cross⍰cohort) design applying multistage sampling strategies. In MoMo 2.0, sociodemographic and anthropometric data of 3,978 individuals (supplement, Table S1) were collected in the field according to a standardized protocol.(13)

### Measures

Demographic variables (e.g., age, sex) were reported by parents for children younger than 11 years and self-reported by adolescents aged 11-17 years. Weight status was based on body mass index (BMI). BMI was computed from individual body weight and height measured by test instructors using a digital scale (Seca 813) and a stadiometer (Seca 213). Overweight was defined as a BMI□>□90th percentile; obesity as a BMI□>□97th percentile, based on the German national reference system.(14)

### Statistical Analytic Strategy

To enable valid comparisons across the two studies over the different assessment waves, we applied an alignment strategy that accounted for differences in study design, sampling procedures, selection probabilities, and non-response rates across KiGGS, MoMo, and MoMo 2.0. MoMo and MoMo 2.0 samples were aligned with data from KiGGS baseline and wave 2. All analyses were conducted in R. The R Package *childsds* (15) was used to calculate BMI and BMI percentiles while the *survey* package (16) was used to calculate weighted prevalence estimates. The analytic code used to implement the alignment strategy and estimate the prevalence rates is publicly available on GitHub at https://github.com/lazaricl/Youth-Overweight-and-Obesity-Germany-KiGGS-MoMo.

#### Sampling Weights

Sampling weights were constructed in two sequential steps. First, design weights were calculated as the inverse of each respondent’s inclusion probability. Second, calibration weights were applied to align the weighted sample distributions of key sociodemographic characteristics including age, sex, education, federal state, and community size classification with known population totals from the microcensus 2023.(17) The final sampling weight, used in all estimations, is the product of the design and calibration weight.(18) The sample weights used in this analysis were trimmed at the upper and lower 2.5% of the original distribution, normalized for all participants excluding participants who completed an online questionnaire only.

#### Alignment

To achieve aligned samples across study designs and sampling strategy, a logistic regression model was fitted using the KiGGS baseline sample predicting MoMo participation. The model included interaction terms of KiGGS BMI * sex, and KiGGS BMI * age. For each subject, the predicted probability of MoMo participation was extracted from the fitted model and the inverse probability of participation weight (IPPW) was obtained.(9, 10) These IPPWs were combined with the sampling weights of each wave within each study. The resulting composite weights were used to calculate aligned, weighted prevalence estimates for overweight and obesity for MoMo baseline and wave 2, and for MoMo 2.0. Subsequently aligned and weighted prevalence estimates were calculated for subgroups (sex, age group, age group * sex). To evaluate this alignment procedure, prevalence rates were compared between MoMo and KiGGS within the corresponding assessment waves. The complete results can be found in the supplement (Table S2-S3).

## Results

### Current Prevalence of Overweight and Obesity (MoMo 2.0)

During 2023-2025, overweight among German youth aged□4-17 years was 16.1% (95% CI [14.5-17.9%]), and obesity prevalence was 6.7% (95% CI [5.5-8.1%], S2-S3). Overweight and obesity prevalence increased with age. The overweight and obesity estimates did not differ between males and females. The changes in excess weight across age groups did not differ by sex (Figure 1).

**Figure 1:**
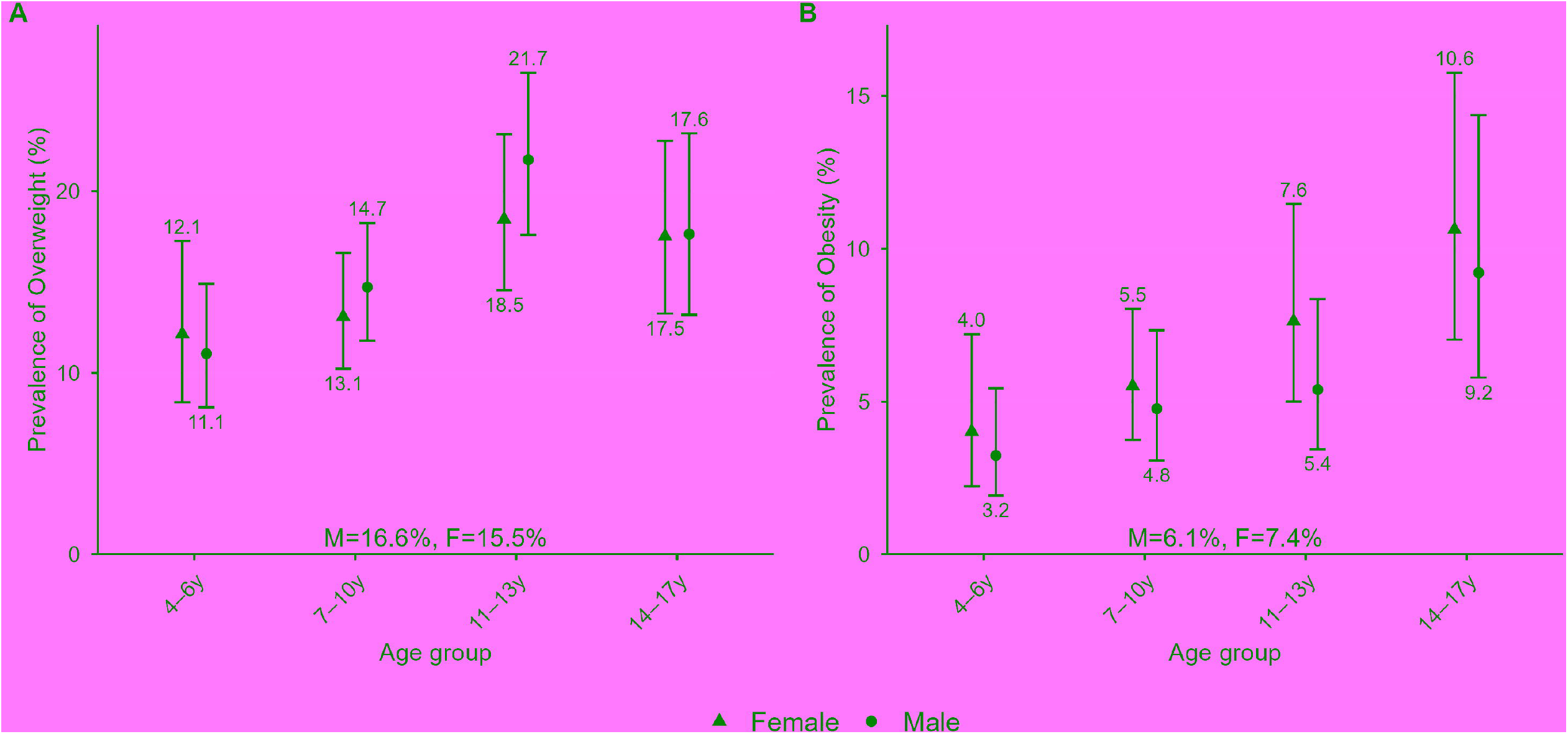
Prevalence of Overweight (A) and Obesity (B) by Sex across Age Groups in MoMo2.0. Circles indicate estimates for males, triangles for females. Error bars show 95% confidence intervals (CI). Y-axes are adapted to the value range of the estimates, the upper and lower CI bounds within each subgroup to avoid overlapping. Rates displayed on top of the x-axes indicate prevalence estimates across all age groups for males (M) and females (F) separately. Numbers displayed above and below the error bars indicate point estimates of specific subgroups (e.g., 4-6y, males). All results are displayed in Tables S2-S3 (supplement).

### Temporal Trend of Overweight and Obesity

In MoMo 2.0, prevalence estimates aligned closely with those from previous KiGGS waves. The overweight prevalence in KiGGS baseline was 16.3% (95% CI [15.4-17.2%]; 2003-2006), and remained stable across wave 2 (16.3%, 95% CI [14.5-18.3%]; 2014-2017), and MoMo 2.0 (16.1% (95% CI [14.5-17.9%]; 2023-2025, Table 1). The obesity prevalence in KiGGS baseline was 7.4% (95% CI [6.8-8.0%]), in wave 2 6.4% (95% CI [5.4-7.6]), and 6.7% in MoMo 2.0 (95% CI [5.5-8.1], Table S2-S3, Figure S1).

**Table 1:**
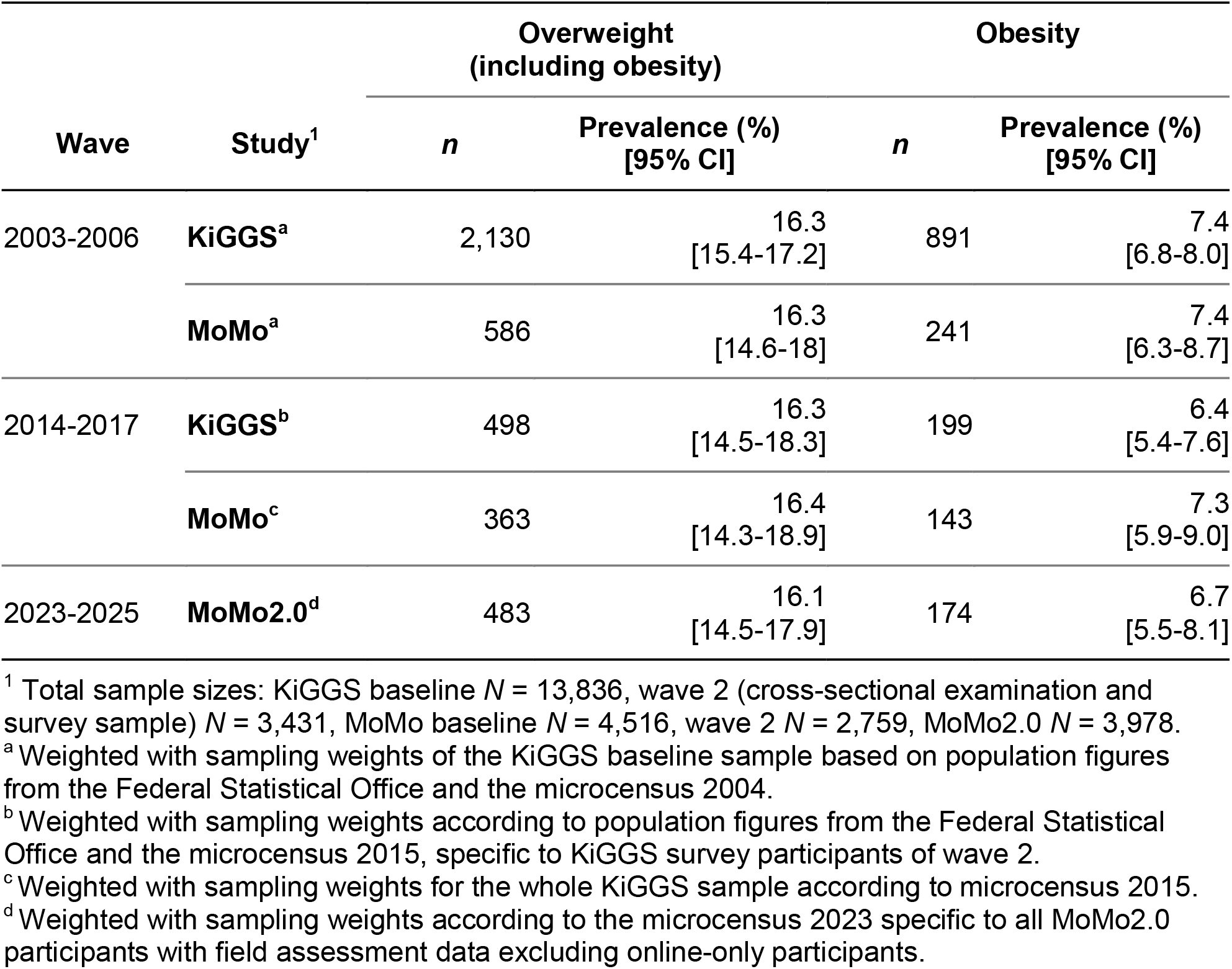
Temporal Trend of Overall Prevalence Estimates of Overweight and Obesity Across the Three Assessment Waves in KiGGS, MoMo and MoMo2.0.

| Wave | Study<sup>1</sup> | Overweight<br>(including obesity) |  | Obesity |  |
| --- | --- | --- | --- | --- | --- |
|  |  | *n* | Prevalence (%)<br>[95% CI] | *n* | Prevalence (%)<br>[95% CI] |
| 2003-2006 | KiGGS<sup>a</sup> | 2,130 | 16.3<br>[15.4-17.2] | 891 | 7.4<br>[6.8-8.0] |
|  | MoMo<sup>a</sup> | 586 | 16.3<br>[14.6-18] | 241 | 7.4<br>[6.3-8.7] |
| 2014-2017 | KiGGS<sup>b</sup> | 498 | 16.3<br>[14.5-18.3] | 199 | 6.4<br>[5.4-7.6] |
|  | MoMo<sup>c</sup> | 363 | 16.4<br>[14.3-18.9] | 143 | 7.3<br>[5.9-9.0] |
| 2023-2025 | MoMo2.0<sup>d</sup> | 483 | 16.1<br>[14.5-17.9] | 174 | 6.7<br>[5.5-8.1] |

<sup>1</sup> Total sample sizes: KiGGS baseline *N* = 13,836, wave 2 (cross-sectional examination and survey sample) *N* = 3,431, MoMo baseline *N* = 4,516, wave 2 *N* = 2,759, MoMo2.0 *N* = 3,978.
<sup>a</sup> Weighted with sampling weights of the KiGGS baseline sample based on population figures from the Federal Statistical Office and the microcensus 2004.
<sup>b</sup> Weighted with sampling weights according to population figures from the Federal Statistical Office and the microcensus 2015, specific to KiGGS survey participants of wave 2.
<sup>c</sup> Weighted with sampling weights for the whole KiGGS sample according to microcensus 2015.
<sup>d</sup> Weighted with sampling weights according to the microcensus 2023 specific to all MoMo2.0 participants with field assessment data excluding online-only participants.

## Discussion

The present study provides the most recent and most reliable population-representative prevalence estimates of overweight and obesity in children and adolescents aged 4-17 years living in Germany. The aligned prevalence of overweight was 16.1%, and 6.7% for obesity. Overweight prevalence increased with age, peaking in early-to-mid adolescence before stabilizing in late adolescence. Obesity prevalence increased with advancing age from 4-17 years across males and females. Sex differences were modest overall.

Based on these study results, overweight and obesity in German youth remained remarkably stable during the last decade. Similar stabilization has been reported across Europe, albeit with considerable cross-national variation (19), whereas global overweight prevalence continues to rise, particularly in high-income countries.(20) Notably, the increases in overweight reported during the COVID-19-pandemic did not continue according to the current estimates. This stabilization may suggest some effect of public health efforts in halting a previously observed upward trend; however, without systematic evaluation of specific prevention strategies, such causal inference is not possible. Given well-documented links between childhood overweight and chronic disease across the life course (3), the resulting public health burden remains substantial.

Since KiGGS and MoMo use partially distinct sampling approaches, IPPW was applied to align samples and enable valid trend comparisons in the absence of current KiGGS data. This newly developed alignment approach, with publicly available analysis scripts on GitHub, represents a methodological strength and may facilitate future population-based research. Nevertheless, the validity of prevalence estimates for small subgroups remains a concern. While this approach works well for the reported estimates, comparability of prevalence rates in small subgroups may be limited. Additionally, the applicability of Kromeyer-Hauschild reference norms to more recent, ethnically diverse populations may be limited.

The current study shows that according to the national reference system roughly one in six children is affected by overweight and nearly one in fourteen by obesity—despite an increased awareness for the harmful effects of excess weight in early life. Structural and behavioral change interventions with demonstrated population-level effectiveness are still to be implemented in Germany, including restrictions on food marketing to children, systemic promotion of physical activity across all settings, access to affordable healthy food and physical activity, and health-promoting school and neighborhood environments. Overweight and obesity in youth remain an unresolved public health challenge, underscoring the need to evaluate existing policies and to develop targeted, evidence-based interventions for high-risk subpopulations. Future research should examine differential trends across socioeconomic subgroups, where burden may be disproportionately high, and move beyond weight status toward multi-metric assessments of body composition. Regular, aligned surveillance across different studies like KiGGS and MoMo 2.0 will be essential to determine whether the current plateau persists or reverses.

## Data Availability

The analytic code used to implement the alignment strategy and estimate the prevalence rates is publicly available on GitHub at https://github.com/lazaricl/Youth-Overweight-and-Obesity-Germany-KiGGS-MoMo.

## REFERENCES

1. Zhang x, Liu J, Ni Y, Yi C, Fang Y, Ning Q, et al. Global Prevalence of Overweight and Obesity in Children and Adolescents: A Systematic Review and Meta-Analysis. JAMA Pediatr 2024;178: 800–813, °10.1001/jamapediatrics.2024.1576.

2. Poveda NE, Elliott MR, Mehta NK, Cunningham SA. Obesity from Childhood to Mid-adulthood in the United States: A Synthetic Cohort Approach to Measuring Health Trajectories. Epidemiology 2026;37: 121–131, °10.1097/EDE.0000000000001927.

3. Twig G, Yaniv G, Levine H, Leiba A, Goldberger N, Derazne E, et al. Body-Mass Index in 2.3 Million Adolescents and Cardiovascular Death in Adulthood. N Engl J Med 2016;374: 2430–2440, °10.1056/NEJMoa1503840.

4. Gallagher C, Pirkis J, Lambert KA, Perret JL, Ali GB, Lodge CJ, et al. Life course BMI trajectories from childhood to mid-adulthood are differentially associated with anxiety and depression outcomes in middle age. Int J Obes (Lond) 2023;47: 661–668, °10.1038/s41366-023-01312-6.

5. Ling J, Chen S, Zahry NR, Kao TA. Economic burden of childhood overweight and obesity: A systematic review and meta-analysis. Obes Rev 2023;24: e13535. °10.1111/obr.13535.

6. Motevalli M, Drenowatz C, Tanous DR, Khan NA, Wirnitzer K. Management of Childhood Obesity-Time to Shift from Generalized to Personalized Intervention Strategies. Nutrients 2021;13, °10.3390/nu13041200.

7. Europe WROf. WHO European Regional Obesity Report 2022. Copenhagen, 2022,°Licence: CC BY-NC-SA 3.0 IGO.

8. Schienkiewitz A, Brettschneider A-K, Damerow S, Schaffrath Rosario A. Übergewicht und Adipositas im Kindes-und Jugendalter in Deutschland – Querschnittergebnisse aus KiGGS Welle 2 und Trends. Journal of Health Monitoring 2018;3, °10.17886/RKI-GBE-2018-005.2.

9. Austin PC, Stuart EA. Moving towards best practice when using inverse probability of treatment weighting (IPTW) using the propensity score to estimate causal treatment effects in observational studies. Stat Med 2015;34: 3661–3679, °10.1002/sim.6607.

10. Westreich D, Edwards JK, Lesko CR, Stuart E, Cole SR. Transportability of Trial Results Using Inverse Odds of Sampling Weights. Am J Epidemiol 2017;186: 1010–1014, °10.1093/aje/kwx164.

11. Mauz E, Lange M, Houben R, Hoffmann R, Allen J, Gosswald A, et al. Cohort profile: KiGGS cohort longitudinal study on the health of children, adolescents and young adults in Germany. Int J Epidemiol 2020;49: 375–375k, °10.1093/ije/dyz231.

12. Woll A, Klos L, Worth A, Hanssen-Doose A, Hinz T, Volkle M, et al. The German national cohort study on the development of motor performance, physical activity and health in children and adolescents: the MoMo 2.0-Study protocol. BMJ Open 2025;15: e094895. °10.1136/bmjopen-2024-094895.

13. Worth A, Hanssen-Doose A, Heinisch S, Nessner C, Opper E, Bös K, et al. MoMo 2.0: Nationale Studie zur Entwicklung von motorischer Leistungsfähigkeit, körperlich-sportlicher Aktivität und Gesundheit von Kindern und Jugendlichen vor dem Hintergrund gesellschaftlicher Veränderungen. Aktualisiertes Testmanual zu den motorischen Tests und den anthropometrischen Messungen der MoMo 2.0-Studie (KIT Scientific Reports, 7760). In: Woll A (ed). KIT Scientific Publishing, 2025,°10.5445/KSP/1000135470.

14. Kromeyer-Hauschild K, Wabitsch M, Kunze D, Geller F, Geiß HC, Hesse V, et al. Perzentile für den Body-mass-Index für das Kindes-und Jugendalter unter Heranziehung verschiedener deutscher Stichproben. Monatsschrift Kinderheilkunde 2001;149: 807–818, °10.1007/s001120170107.

15. Vogel M. childsds: Data and Methods Around Reference Values in Pediatrics. 2025

16. Lumley T. survey: Analysis of Complex Survey Samples. 2026

17. Statistisches Bundesamt D. Qualitätsbericht Mikrozensus 20222. Statistisches Bundesamt (Destatis) 2025.

18. Sand M, Kunz T. Gewichtung in der Praxis (Version 1.0) [GESIS Survey Guidelines]. Mannheim: GESIS Leibniz-Institut für Sozialwissenschaften 2020, °10.15465/gesis-sg_030.

19. Buoncristiano M, Spinelli A, Williams J, Nardone P, Rito AI, Garcia-Solano M, et al. Childhood overweight and obesity in Europe: Changes from 2007 to 2017. Obes Rev 2021;22 Suppl 6: e13226. °10.1111/obr.13226.

20. Collaborators GAB. Global, regional, and national prevalence of child and adolescent overweight and obesity, 1990-2021, with forecasts to 2050: a forecasting study for the Global Burden of Disease Study 2021. Lancet 2025;405: 785–812, °10.1016/S0140-6736(25)00397-6.

