## Supplement file for "Population-Based Prevalence of Overweight and Obesity in German Youth: Findings from MoMo 2.0 and Comparison with KiGGS"

Claudia Lazarides
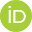
<https://orcid.org/0000-0002-8249-3560>^1^, Ronny Kuhnert^2^, Anja Schienkiewitz^2^, Lara Tschuschke
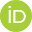
<https://orcid.org/0009-0004-0068-6580>^3^, Leon Klos
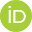
<https://orcid.org/0000-0001-8236-1309>^3^, Janis Fiedler^3^, Laura Wolbring^3^, Anke Hanssen-Doose^4^, Carmen Volk
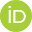
<https://orcid.org/0000-0002-8480-9514>^3^, Claudia Niessner^3^, Julika Loss^2^, Manuel C. Voelkle
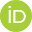
<https://orcid.org/0000-0001-5576-8103>^1^ & Alexander Woll^3^

^1^ Department of Psychology, Division Evaluation, University of Freiburg, Freiburg im Breisgau, Germany.

*Address:* Department of Psychology, Division Evaluation, University of Freiburg, Engelbergerstraße 41, 79085 Freiburg im Breisgau, Germany

^2^ Department of Epidemiology and Health Monitoring, Robert Koch Institute, Berlin, Germany.

*Address:* Department of Epidemiology and Health Monitoring, Robert Koch Institute, Gerichtstraße 27, 13347 Berlin, Germany

^3^ Institute of Sports and Sports Science, Karlsruhe Institute of Technology, Karlsruhe, Germany.

*Address:* Institute of Sports and Sports Science, Karlsruhe Institute of Technology (KIT), Engler-Bunte-Ring 15, 76131 Karlsruhe, Germany

^4^ Institute of Movement and Sport, Karlsruhe University of Education, Karlsruhe, Germany.

*Address:* Institute of Movement and Sport, Karlsruhe University of Education, Bismarckstraße 10, 76133 Karlsruhe, Germany

**Corresponding Author:** Claudia Lazarides, Department of Psychology, University of Freiburg, 79106 Freiburg, Germany.

**Information on co-authors**

| **Name** | **Surname** | **E-Mail** |
| --- | --- | --- |
| Claudia | Lazarides | |
| Ronny | Kuhnert | |
| Anja | Schienkiewitz | |
| Lara | Tschuschke | |
| Leon | Klos | |
| Janis | Fiedler | |
| Laura | Wolbring | |
| Anke | Hanssen-Doose | |
| Carmen | Volk | |
| Claudia | Niessner | |
| Julika | Loss | |
| Manuel C. | Voelkle | |
| Alexander | Woll | |

**Table S1:** Sample description of MoMo 2.0: Sociodemographic and anthropometric characteristics by sex.

| **Variable** | **Overall**  N = 3,967^1,2^ | **Male**  n = 2,078^2^ | **Female**  n = 1,889^2^ |
| --- | --- | --- | --- |
| **Age** | 10.42 (3.71) | 10.42 (3.66) | 10.41 (3.76) |
| **Age groups** |  |  |  |
| 4–6y | 576 (15%) | 296 (14%) | 280 (15%) |
| 7–10y | 1,677 (42%) | 875 (42%) | 802 (42%) |
| 11–13y | 912 (23%) | 491 (24%) | 421 (22%) |
| 14–17y | 802 (20%) | 416 (20%) | 386 (20%) |
| **SES category** |  |  |  |
| Low | 743 (19%) | 395 (20%) | 348 (19%) |
| Medium | 2,316 (61%) | 1,208 (61%) | 1,108 (61%) |
| High | 754 (20%) | 391 (20%) | 363 (20%) |
| Unknown | 154 | 84 | 70 |
| **Migration background** | |  |  |
| None | 2,784 (73%) | 1,445 (72%) | 1,339 (73%) |
| One-sided | 451 (12%) | 236 (12%) | 215 (12%) |
| Two-sided | 595 (16%) | 326 (16%) | 269 (15%) |
| Unknown | 137 | 71 | 66 |
| **Height (cm)** | 143.05 (21.77) | 144.46 (22.82) | 141.48 (20.44) |
| **Weight (kg)** | 39.34 (17.67) | 40.23 (18.60) | 38.35 (16.53) |
| **BMI^3^ category** |  |  |  |
| Obesity (>P97) | 174 (4.4%) | 86 (4.1%) | 88 (4.7%) |
| Overweight (>P90–P97) | 309 (7.8%) | 168 (8.1%) | 141 (7.5%) |
| Normal weight | 3,186 (80%) | 1,676 (81%) | 1,510 (80%) |
| Underweight^4^ (P3 –<P10) | 212 (5.3%) | 99 (4.8%) | 113 (6.0%) |
| Severe underweight^4^ (<P3) | 86 (2.2%) | 49 (2.4%) | 37 (2.0%) |
| *Note.* Displayed are unweighted descriptive statistics.  ^1^ *n* = 2 diverse participants were excluded due to missing BMI norms and to guarantee anonymity. *n* = 9 participants aged older than 18 at assessment were excluded due to missing BMI norms.  ^2^ Mean (SD); n (%)  ^3^ BMI = Body Mass Index based on Kromeyer-Hauschild norms (2001).  ^4^ Weight status is also referred to as *thinness* in international reference systems. | | | |

**Table S2:** Prevalence of Overweight Including Obesity (BMI>90th Percentile) with 95% Confidence Interval for Total Sample, by Sex, by Age Group, and by Sex and Age Group for Study Waves 2003-2006, and 2014-2017 in KiGGS, and MoMo, and 2023-2025 in MoMo 2.0 with and without Alignment.

|  |  | **KiGGS** | | | | **MoMo/MoMo 2.0** | | | | | | **Aligned MoMo/MoMo 2.0** | | | | | |
| --- | --- | --- | --- | --- | --- | --- | --- | --- | --- | --- | --- | --- | --- | --- | --- | --- | --- |
|  |  | **2003-2006^a^** | | **2014-2017^b^** | | **2003-2006^a^** | | **2014-2017^c^** | | **2023-2025^d^** | | **2003-2006^a^** | | **2014-2017^c^** | | **2023-2025^d^** | |
|  |  | **Prevalence^1^**  **[95% CI]** | ***n*** | **Prevalence^1^**  **[95% CI]** | ***n*** | **Prevalence^1^**  **[95% CI]** | ***n*** | **Prevalence^1^**  **[95% CI]** | ***n*** | **Prevalence^1^**  **[95% CI]** | ***n*** | **Prevalence^1^**  **[95% CI]** | ***n*** | **Prevalence^1^**  **[95% CI]** | ***n*** | **Prevalence^1^**  **[95% CI]** | ***n*** |
| **Total sample** | | 16.3 [15.4-17.2] | 2,130 | 16.3 [14.5-18.3] | 498 | 13.5 [12.1-15.0] | 586 | 13.1 [11.4-14.9] | 363 | 12.8 [11.6-14.2] | 483 | 16.3 [14.6-18.0] | 586 | 16.4 [14.3-18.9] | 363 | 16.1 [14.5-17.9] | 483 |
| **Sex** | **Age Group** |  |  |  |  |  |  |  |  |  |  |  |  |  |  |  |  |
| M |  | 16.2 [15.0-17.5] | 1,096 | 16.1 [13.3-19.2] | 242 | 12.8 [11.0-14.7] | 293 | 13.2 [11.0-15.8] | 178 | 12.8 [11.2-14.7] | 254 | 16.0 [13.9-18.4] | 293 | 17.7 [14.6-21.2] | 178 | 16.6 [14.4-19.2] | 254 |
| F |  | 16.3 [15.2-17.4] | 1,034 | 16.5 [14.3-19.1] | 256 | 14.3 [12.6-16.1] | 293 | 13.0 [10.8-15.5] | 185 | 12.9 [11.0-14.9] | 229 | 16.5 [14.6-18.7] | 293 | 15.1 [12.4-18.2] | 185 | 15.5 [13.3-17.9] | 229 |
|  | 4-6y | 9.6 [8.2-11.2] | 274 | 9.9 [7.1-13.6] | 58 | 8.9 [7.2-11.0] | 133 | 7.7 [5.4-10.7] | 47 | 9.0 [6.8-11.8] | 50 | 9.5 [7.6-11.9] | 133 | 8.7 [6.3-12.0] | 47 | 11.2 [8.4-14.7] | 50 |
|  | 7-10y | 16.2 [14.9-17.5] | 652 | 15.6 [12.6-19.2] | 125 | 14.2 [11.9-16.9] | 166 | 12.4 [9.9-15.4] | 106 | 12.1 [10.3-14.1] | 183 | 16.2 [13.6-19.2] | 166 | 14.5 [11.4-18.2] | 106 | 13.7 [11.7-15.9] | 183 |
|  | 11-13y | 19.4 [17.6-21.3] | 569 | 20.2 [16.4-24.5] | 140 | 16.7 [13.2-20.9] | 138 | 17.9 [14.0-22.6] | 108 | 17.1 [14.7-19.8] | 148 | 19.4 [15.4-24.1] | 138 | 22.2 [17.3-28.0] | 108 | 20.2 [17.4-23.4] | 148 |
|  | 14-17y | 18.3 [16.9-19.8] | 635 | 18.3 [15.7-21.4] | 175 | 15.9 [13.3-18.9] | 149 | 12.9 [10.3-16.1] | 102 | 13.4 [11.1-16.0] | 102 | 18.4 [15.4-21.7] | 149 | 16.3 [12.8-20.4] | 102 | 17.6 [14.5-21.2] | 102 |
| M | 4-6y | 9.6 [7.7-11.9] | 133 | 8.0 [4.9-12.8] | 28 | 8.3 [6.0-11.5] | 61 | 7.1 [4.5-11.2] | 22 | 8.1 [5.5-11.7] | 25 | 9.4 [6.6-13.1] | 61 | 8.5 [5.4-13.1] | 22 | 10.0 [6.8-14.7] | 25 |
| M | 7-10y | 16.8 [14.9-18.8] | 348 | 16.1 [11.6-21.7] | 66 | 14.1 [10.7-18.3] | 87 | 11.6 [7.8-16.9] | 47 | 12.3 [10.0-15.0] | 95 | 16.9 [12.9-21.7] | 87 | 14.5 [9.6-21.1] | 47 | 14.4 [11.8-17.5] | 95 |
| M | 11-13y | 18.6 [16.4-21.1] | 286 | 20.6 [15.1-27.5] | 70 | 14.5 [10.8-19.2] | 69 | 19.4 [14.6-25.4] | 58 | 17.4 [14.0-21.4] | 82 | 18.1 [13.7-23.4] | 69 | 25.6 [19.3-33.2] | 58 | 21.7 [17.6-26.5] | 82 |
| M | 14-17y | 18.2 [16.3-20.3] | 329 | 18.1 [13.9-23.2] | 78 | 15.5 [12.0-19.9] | 76 | 12.8 [9.2-17.5] | 51 | 13.2 [10.0-17.2] | 52 | 18.0 [13.9-22.9] | 76 | 17.0 [12.1-23.2] | 51 | 17.6 [13.2-23.2] | 52 |
| F | 4-6y | 9.6 [7.9-11.6] | 141 | 11.9 [7.2-18.9] | 30 | 9.5 [7.2-12.5] | 72 | 8.2 [5.0-13.1] | 25 | 9.9 [6.7-14.4] | 25 | 9.7 [7.3-12.7] | 72 | 8.9 [5.6-13.9] | 25 | 12.3 [8.4-17.7] | 25 |
| F | 7-10y | 15.6 [13.7-17.6] | 304 | 15.1 [11.0-20.5] | 59 | 14.4 [11.3-18.2] | 79 | 13.2 [9.8-17.6] | 59 | 11.9 [9.5-14.8] | 88 | 15.5 [12.3-19.5] | 79 | 14.5 [10.7-19.2] | 59 | 12.9 [10.3-15.9] | 88 |
| F | 11-13y | 20.1 [17.7-22.8] | 283 | 19.7 [14.7-25.8] | 70 | 19.1 [14.4-25.0] | 69 | 16.1 [11.2-22.8] | 50 | 16.8 [13.2-21.1] | 66 | 20.9 [15.7-27.2] | 69 | 18.0 [12.3-25.6] | 50 | 18.5 [14.5-23.1] | 66 |
| F | 14-17y | 18.4 [16.3-20.7] | 306 | 18.6 [14.7-23.2] | 97 | 16.2 [12.4-20.8] | 73 | 13.1 [9.3-18.1] | 51 | 13.5 [10.4-17.4] | 50 | 18.7 [14.5-23.9] | 73 | 15.4 [10.9-21.5] | 51 | 17.5 [13.3-22.8] | 50 |
| *Note.* M = Male, F = Female. *n* = size of subgroup. Total sample size: KiGGS baseline *N* = 13,836, wave 2 (cross-sectional examination and survey sample) *N* = 3,431, MoMo baseline *N* = 4,516, wave 2 *N* = 2,759, MoMo 2.0 *N* = 3,977.  ^a^ Weighted with sampling weights of the KiGGS baseline sample based on population figures from the Federal Statistical Office and the microcensus 2004.  ^b^ Weighted with sampling weights according to population figures from the Federal Statistical Office and the microcensus 2015, specific to KiGGS survey participants of wave 2.  ^c^ Weighted with sampling weights for the whole KiGGS sample according to population figures from the Federal Statistical Office and to microcensus 2015.  ^d^ Weighted with sampling weights according to the microcensus 2023 specific to all MoMo2.0 participants with field assessment data excluding online-only participants.  ^1^ Prevalence in percent (%) | | | | | | | | | | | | | | | | | |

**Table S3:** Prevalence of Obesity (BMI>97th Percentile) with 95% Confidence Interval for Total Sample, by Sex, by Age Group, and by Sex and Age Group for Study Waves 2003-2006, and 2014-2017 in KiGGS, and in MoMo, and 2023-2025 in MoMo 2.0 with and without Alignment.

|  |  | **KiGGS** | | | | **MoMo/MoMo 2.0** | | | | | | **Aligned MoMo/MoMo 2.0** | | | | | |
| --- | --- | --- | --- | --- | --- | --- | --- | --- | --- | --- | --- | --- | --- | --- | --- | --- | --- |
|  |  | **2003-2006^a^** | | **2014-2017^b^** | | **2003-2006^a^** | | **2014-2017^c^** | | **2023-2025^d^** | | **2003-2006^a^** | | **2014-2017^c^** | | **2023-2025^d^** | |
|  |  | **Prevalence^1^ [95% CI]** | **n** | **Prevalence^1^ [95% CI]** | **n** | **Prevalence^1^ [95% CI]** | **n** | **Prevalence^1^ [95% CI]** | **n** | **Prevalence^1^ [95% CI]** | **n** | **Prevalence^1^ [95% CI]** | **n** | **Prevalence^1^ [95% CI]** | **n** | **Prevalence^1^ [95% CI]** | **n** |
| **Total Sample** | | 7.4 [6.8-8.0] | 891 | 6.4 [5.4-7.6] | 199 | 5.5 [4.7-6.5] | 241 | 5.5 [4.5-6.7] | 143 | 4.9 [4.1-5.9] | 174 | 7.4 [6.3-8.7] | 241 | 7.3 [5.9-9.0] | 143 | 6.7 [5.5-8.1] | 174 |
| **Sex** | **Age Group** |  |  |  |  |  |  |  |  |  |  |  |  |  |  |  |  |
| M |  | 7.2 [6.4-8.1] | 462 | 6.6 [5.1-8.5] | 100 | 5.4 [4.4-6.6] | 127 | 5.7 [4.3-7.6] | 70 | 4.6 [3.6-6.0] | 86 | 6.9 [5.5-8.5] | 127 | 7.8 [5.8-10.4] | 70 | 6.1 [4.6-8.1] | 86 |
| F |  | 7.6 [6.8-8.4] | 429 | 6.2 [4.9-7.9] | 99 | 5.7 [4.6-7.1] | 114 | 5.2 [4.0-6.9] | 73 | 5.3 [4.1-6.7] | 88 | 7.9 [6.4-9.8] | 114 | 6.8 [5.1-9.0] | 73 | 7.4 [5.7-9.5] | 88 |
|  | 4-6y | 3.5 [2.7-4.4] | 101 | 2.1 [1.0-4.1] | 14 | 3.7 [2.6-5.2] | 52 | 2.3 [1.4-3.9] | 17 | 2.6 [1.5-4.4] | 16 | 3.5 [2.4-5.0] | 52 | 2.2 [1.3-3.7] | 17 | 2.2 [1.3-3.8] | 16 |
|  | 7-10y | 7.2 [6.3-8.2] | 273 | 5.8 [4.0-8.3] | 49 | 5.8 [4.4-7.5] | 75 | 5.9 [4.0-8.7] | 42 | 4.5 [3.4-5.9] | 63 | 7.2 [5.6-9.2] | 75 | 7.6 [5.1-11.3] | 42 | 5.3 [4.1-6.9] | 63 |
|  | 11-13y | 8.0 [6.7-9.4] | 209 | 7.3 [5.1-10.4] | 48 | 6.7 [4.7-9.6] | 52 | 6.3 [4.3-9.2] | 36 | 5.5 [4.1-7.4] | 46 | 8.0 [5.6-11.3] | 52 | 7.3 [5.0-10.7] | 36 | 6.4 [4.8-8.5] | 46 |
|  | 14-17y | 9.5 [8.4-10.8] | 308 | 9.1 [6.9-11.8] | 88 | 6.7 [5.0-8.8] | 62 | 6.2 [4.5-8.6] | 48 | 6.8 [5.0-9.2] | 49 | 9.5 [7.2-12.5] | 62 | 8.5 [6.1-11.9] | 48 | 9.9 [7.2-13.5] | 49 |
| M | 4-6y | 2.8 [2.0-4.0] | 50 | 1.1 [0.4-3.2] | 4 | 3.6 [2.2-5.8] | 27 | 1.8 [0.8-4.4] | 7 | 2.5 [1.3-4.9] | 9 | 3.4 [2.1-5.5] | 27 | 1.8 [0.8-4.1] | 7 | 2.2 [1.1-4.2] | 9 |
| M | 7-10y | 7.8 [6.5-9.4] | 155 | 6.8 [4.2-11.0] | 25 | 6.2 [4.4-8.8] | 44 | 5.3 [2.6-10.6] | 16 | 4.1 [2.7-6.2] | 29 | 7.5 [5.3-10.6] | 44 | 6.9 [3.3-14.0] | 16 | 4.9 [3.2-7.3] | 29 |
| M | 11-13y | 7.6 [6.0-9.7] | 105 | 8.1 [5.0-13.1] | 28 | 6.3 [3.7-10.5] | 27 | 6.8 [4.1-11.2] | 18 | 4.8 [3.1-7.5] | 23 | 7.4 [4.4-12.3] | 27 | 7.7 [4.6-12.6] | 18 | 5.4 [3.4-8.3] | 23 |
| M | 14-17y | 9.0 [7.6-10.6] | 152 | 8.8 [5.9-12.8] | 43 | 6.0 [4.0-9.1] | 29 | 7.4 [4.9-11.0] | 29 | 6.6 [4.2-10.1] | 25 | 8.0 [5.3-12.0] | 29 | 10.1 [6.5-15.2] | 29 | 9.2 [5.8-14.4] | 25 |
| F | 4-6y | 4.1 [3.0-5.8] | 51 | 3.0 [1.2-7.2] | 10 | 3.7 [2.3-6.1] | 25 | 2.7 [1.3-5.5] | 10 | 2.6 [1.2-5.7] | 7 | 3.5 [2.1-5.8] | 25 | 2.7 [1.3-5.3] | 10 | 2.3 [1.0-4.9] | 7 |
| F | 7-10y | 6.5 [5.4-7.8] | 118 | 4.7 [2.9-7.5] | 24 | 5.3 [3.5-7.9] | 31 | 6.6 [4.3-10.0] | 26 | 4.8 [3.3-7.0] | 34 | 6.8 [4.6-10.0] | 31 | 8.4 [5.6-12.5] | 26 | 5.9 [4.1-8.2] | 34 |
| F | 11-13y | 8.3 [6.6-10.4] | 104 | 6.5 [3.7-11.2] | 20 | 7.2 [4.4-11.7] | 25 | 5.8 [3.5-9.4] | 18 | 6.4 [4.2-9.6] | 23 | 8.6 [5.3-13.7] | 25 | 6.9 [4.2-11.0] | 18 | 7.6 [5.0-11.5] | 23 |
| F | 14-17y | 10.0 [8.5-11.9] | 156 | 9.4 [6.5-13.4] | 45 | 7.3 [4.9-10.6] | 33 | 4.9 [3.0-8.1] | 19 | 7.0 [4.7-10.3] | 24 | 11.0 [7.5-15.8] | 33 | 6.8 [4.0-11.3] | 19 | 10.6 [7.0-15.8] | 24 |
| *Note.* M = Male, F = Female. *n* = size of subgroup. Total sample size: KiGGS baseline *N* = 13,836, wave 2 (cross-sectional examination and survey sample) *N* = 3,431, MoMo baseline *N* = 4,516, wave 2 *N* = 2,759, MoMo2.0 *N* = 3,977.  ^a^ Weighted with sampling weights of the KiGGS baseline sample based on population figures from the Federal Statistical Office and the microcensus 2004.  ^b^ Weighted with sampling weights according to population figures from the Federal Statistical Office and the microcensus 2015, specific to KiGGS survey participants of wave 2.  ^c^ Weighted with sampling weights for the whole KiGGS sample according to population figures from the Federal Statistical Office and to microcensus 2015.  ^d^ Weighted with sampling weights according to the microcensus 2023 specific to all MoMo2.0 participants with field assessment data excluding online-only participants.  ^1^ Prevalence in percent (%) | | | | | | | | | | | | | | | | | |

**Figure S1:** Temporal Trend of the Prevalence of Overweight (A, B) and Obesity (C, D) by Sex and Age Group Across Three Assessment Waves (2003-2006, 2014-2017, and 2023-2025) of KiGGS, aligned MoMo and MoMo 2.0.
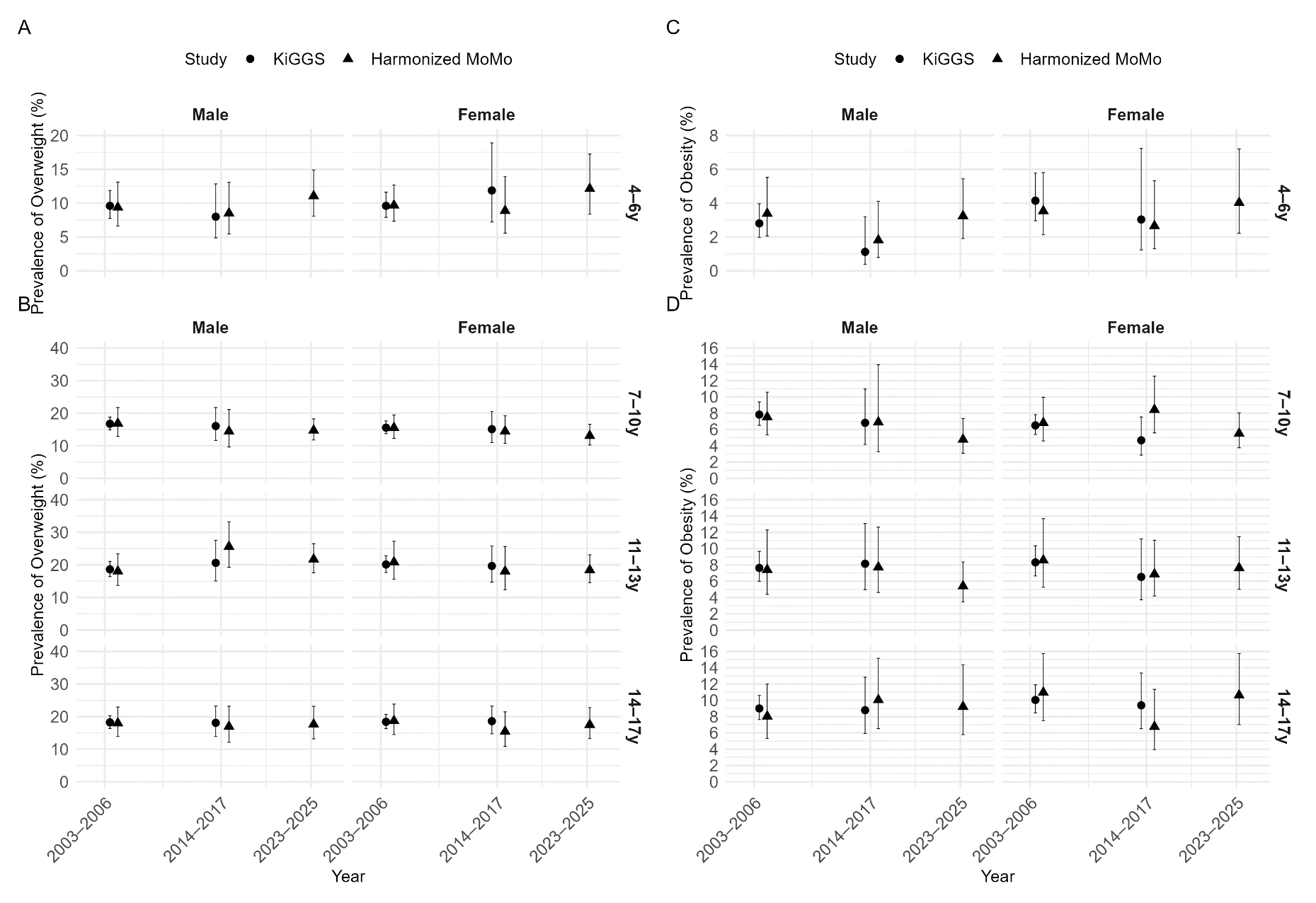

*Caption:* Circles indicate estimates based on KiGGS. Triangles indicate estimates based on MoMo and MoMo 2.0. Error bars show 95 % confidence intervals around the point estimate. Y-axes are adopted to the value range of the estimates, the upper and lower CI bounds within each subgroup to avoid overlap. All results can be found in Table S2-S3.
